# Enhancing the Discriminatory Power of ADHD and Autism Spectrum Disorder Polygenic Scores in Clinical and Non-Clinical Samples

**DOI:** 10.1101/2022.02.09.22270697

**Authors:** James J. Li, Quanfa He, Zihang Wang, Qiongshi Lu

**Affiliations:** Department of Psychology, University of Wisconsin-Madison; Waisman Center, University of Wisconsin-Madison; Center for Demography of Health and Aging, University of Wisconsin-Madison; Department of Statistics, University of Wisconsin-Madison; Department of Biostatistics and Bioinformatics, Emory University; Department of Biostatistics and Medical Informatics, University of Wisconsin-Madison

**Author notes:** Please address all correspondence and reprint requests to James J. Li, Ph.D., Department of Psychology, 1202 W. Johnson Street, Madison, WI 53706. Disclosures James J. Li, Quanfa He, Zihang Wang, and Qiongshi Lu report no financial relationships with commercial interests.

## Abstract

**Objective:** Polygenic scores (PGS) are widely used in psychiatric genetic associations studies due to their impressive power to predict focal outcomes. However, they lack in discriminatory power, in part due to the high degree of genetic overlap between psychiatric disorders. The lack of prediction specificity limits the clinical utility of psychiatric PGS, particularly for diagnostic applications. The goal of the study was to enhance the discriminatory power of psychiatric PGS for two highly comorbid and genetically correlated neurodevelopmental disorders in ADHD and autism spectrum disorder (ASD).

**Methods:** Genomic structural equation modeling (GenomicSEM) was used to generate novel PGS for ADHD and ASD by accounting for the genetic overlap between these disorders (and eight others) to achieve greater discriminatory power in non-focal outcome predictions. PGS associations were tested in two large independent samples – the Philadelphia Neurodevelopmental Cohort (*N*=4,789) and the Simons Foundation Powering Autism Research for Knowledge (SPARK) ASD and sibling controls (*N*=5,045) cohort.

**Results:** PGS from GenomicSEM achieved superior discriminatory power in terms of showing significantly attenuated associations with non-focal outcomes relative to traditionally computed PGS for these disorders. Additionally, genetic correlations between GenomicSEM PGS for ASD and ADHD were significantly attenuated in cross-trait associations with other psychiatric disorders and outcomes.

**Conclusions:** Psychiatric PGS associations are likely inflated by the high degree of genetic overlap between the psychiatric disorders. Methods such as GenomicSEM can be used to refine PGS signals to be more disorder-specific, thereby enhancing their discriminatory power for future diagnostic applications.

---

Psychiatric disorders all have polygenic bases by which numerous DNA variants of individually small effects contribute to their etiologies (1). However, gene identification efforts via genome wide association studies (GWAS) are likely confounded by the fact that psychiatric disorders also feature a high degree of genetic overlap (2). That is, some genes that were previously identified in univariate psychiatric GWAS may not be unique to the disorder, but in fact be contributing to pleiotropy (3,4). Despite this likely confound, single-disorder polygenic scores (PGS) remain widely employed in genetic association studies due to their impressive power to predict focal outcomes. Yet, these advantages are offset by their lack of discriminatory power as they also tend to be associated with a myriad of non-focal outcomes as well. For example, PGS for attention-deficit/hyperactivity disorder (ADHD) are not only predictive of ADHD (5), but they also predict major depression (6), autism spectrum disorder (ASD) (7), substance use disorders (8), and conduct problems (9). Improving the discriminatory power of PGS is critical if they are to be employed in diagnostic applications (10).

This limitation can be addressed by modeling the genetic components of a single disorder via GWAS summary statistics simultaneously with other psychiatric disorders to produce a genetic covariance matrix, using an approach called Genomic Structural Equation Modeling (GenomicSEM) (3). Phenotypically, this model is referred to as the Hierarchical Taxonomy of Psychopathology (HiTOP) (11), in which distinct higher order dimensions capture the etiological commonalities between discrete disorders that tend to cluster (i.e., correlate) together. These include internalizing (e.g., depression, anxiety), externalizing (e.g., attention and behavioral disorders), thought (e.g., schizophrenia, bipolar disorder), and neurodevelopmental (e.g., ASD, ADHD) dimensions. Genetically-informed studies using GWAS summary statistics have uncovered a similar hierarchical structure (2,3,12). With this approach, genetically unique components (i.e., residuals) can be estimated separately from the genetic components that contribute to pleiotropy. PGS can then be weighted by the residual genetic components which are *disorder specific* and thus, more discriminative in their predictive performance relative to traditionally computed PGS.

For this investigation, we focused on enhancing the discriminatory power of PGS for ADHD and ASD specifically. ADHD and ASD are classified as neurodevelopmental disorders (13), where ADHD is characterized by deficits in the inattention and/or hyperactivity/impulsivity domains, and ASD is characterized by deficits in social communication and interaction skills and impairing restrictive or repetitive behaviors. These disorders are challenging to study in tandem, as not only do their clinical presentations overlap considerably, resulting in a high rate of co-morbidity (14), but they are also genetically correlated with one another (15). ADHD and ASD PGS are also associated with a myriad of other phenotypes, and thus should provide a proof-of-concept for the utility of GenomicSEM to enhance discriminatory power for other psychiatric PGS more broadly. Finally, there is a critical need to identify differentiated risk factors for ADHD and ASD given that the appropriate early intervention for this population depends on a precise diagnosis.

## Method

### GWAS Summary Statistics

In order to identify the genomic structure for multiple psychiatric disorders, we used GWAS summary statistics for 10 disorders via the Psychiatric Genomics Consortium and other genetic repositories: ADHD (16), anorexia (AN) (17), anxiety disorders (ANX) (18,19), ASD (15), bipolar disorder (BIP) (20), major depressive disorder (MDD) (21), obsessive compulsive disorder (OCD) (22), post-traumatic stress disorder (PTSD) (23,24), schizophrenia (SCZ) (25), and Tourette syndrome (TS) (26). Details for each GWAS are provided in Table S1. The current investigation prioritized GWAS for those of European ancestry, given that most discovery samples are based on individuals of European ancestry and concerns have been raised about the generalizability of PGS associations in non-European ancestries (10). Following precedent (4), in cases where more than one GWAS was conducted for a disorder, a meta-analysis of the summary statistics was conducted in GenomicSEM while also accounting for overlapping samples.

### Target Samples

Two independent samples were used to examine associations between PGS and focal (ADHD and ASD) versus non-focal (non-ADHD or ASD) outcomes. *The Philadelphia Neurodevelopmental Cohort (PNC)* is a population-based dataset of children, adolescents, and young adults (ages 8-21) with data on psychiatric disorders, medical history, neuroimaging, genetics, and neurocognition. Participants were recruited in the greater Philadelphia, Pennsylvania area between November 2009 to December 2011. Additional information of this study are detailed elsewhere (27). Individuals of European ancestry with both genotypic and phenotypic data were included in the analyses (*N*=4,789). Psychiatric disorders were assessed using a semi-structured, computerized clinical interview adapted from the Kiddie Schedule for Affective Disorders and Schizophrenia (K-SADS). The interview was administered to the caregivers or legal guardians (i.e., collaterals) of participants aged 8 to 10, participants and collaterals who were ages 11 to 17, and the participants themselves if they were between the ages of 18 to 21. To minimize reporter inconsistency between the age groups, we used data from collaterals for participants between 8 to 17 years of age, and from self-report for those older than 18. For our PGS analyses, we analyzed the number of symptoms for the following disorders: MDD, manic episodes (MAN), generalized anxiety disorder (GAD), social anxiety disorder (SOC), separation anxiety disorder (SEP), specific phobias (PHB), agoraphobia (AGR), panic disorder (PAN), AN, ADHD, oppositional defiant disorder (ODD), conduct disorder (CDD), OCD, and psychosis (PSY) (Table S2).

*The Simons Foundation Powering Autism Research for Knowledge (SPARK)* is comprised of individuals with a diagnosis of ASD, their biological parents, and an unaffected sibling from 31 sites in the United States. A total of 251,082 individuals were recruited, including 84,005 ASD individuals under the age of 18. Additional information for this study is available here (28). Two subgroups were examined for our analyses – ASD probands and sibling controls. ASD probands were individuals under the age of 18 with a diagnosis of ASD that also had no affected siblings in the sample (*n*=3,248, mean age=8.27, S.D.=4.05). Controls were under the age of 18 without an ASD diagnosis and had a sibling who was enrolled as an ASD proband (*n*=1,797, mean age =8.10, S.D.=4.50) (see Tables S3-S4). These restrictions accounted for the impact of genetic relatedness among individuals in our analyses. Psychiatric disorders were assessed via a medical screening questionnaire, including for ADHD, ODD, CDD, ANX, BIP, MDD, OCD, SCZ, learning disability (LD), intellectual disability (ID) and TS. ASD outcomes were assessed via parent-reported measures for the two dimensions of ASD: the Repetitive Behavior Scale-Revised (RBSR) (29) and the Social Communication Questionnaire (SCQ) (29). Total scores for the RBSR and SCQ were treated as separate outcomes.

### GenomicSEM and PGS Computations

GenomicSEM calculates the genetic covariance among multiple disorders using GWAS summary statistics, and accounts for any sample overlap across studies via linkage disequilibrium (LD) score regression (30). To identify an optimal GenomicSEM model, we first performed an exploratory factor analysis (EFA) with each available GWAS summary statistic.

The genetic covariance matrix was calculated using LD score regression and was used as the input for EFA. EFA models ranged from 2-5 factors with oblique rotations. Following the identification of an optimal EFA factor structure (based on eigenvalues >1), we specified confirmatory factor analysis (CFA) models based on the EFA loading patterns and assessed model fit using standard fit indices. After an optimal CFA GenomicSEM model was identified, we then estimated each SNP’s association with ADHD and ASD simultaneously, independent of its associations with the latent factors (i.e., adding “SNP edges” to the GenomicSEM model). This model tested for three additional paths of association for each SNP, where SNP associations are generated for each estimated path.

Two versions of ADHD and ASD PGS were computed in our association analyses: 1) “traditional” PGS, which used univariate summary statistics for ADHD and ASD respectively, and 2) “disorder-specific” PGS, computed using summary statistics from our GenomicSEM analysis. A third PGS was computed from GenomicSEM genetic effects estimated for the latent genetic dimension that included ASD and ADHD. In sum, we generated a total of five separate PGS. We used PLINK (31) to clump the GWAS summary statistics using the CEU samples in 1000 Genome Project Phase III cohort (32) as the LD reference panel. We then specified an LD window size of 1000 kb and a LD threshold of 0.1 for clumping. PGS association analyses covaried for biological sex, age, and the first 10 genetic principal components (PCs). All PGS were standardized to a mean of 0 and a variance of 1.

## Results

### Model Fitting and GenomicSEM

Factor loadings and eigenvalues for the EFA 2-5 factor genetic models are provided in Tables S5-S8. CFA models were tested based on what was observed in the EFA factor loadings, thus allowing the data to guide our final model selection in GenomicSEM. CFA models were specified where EFA loadings onto factor dimensions were above .30. For disorders that had no EFA loadings above .30, we specified the disorder to load onto the factor with the highest loading. Five different CFA models were tested, ranging from a unidimensional to a 5-factor model (Table S9). The 3-factor model exhibited the best fit among the five models (AIC=343.091, CFI=.935, SRMR=.101), and the loadings were interpretable. Following empirical precedent (2), and further supported by findings from our EFA model, the optimal GenomicSEM model was one in which ANX, MDD, PTSD, and AN loaded on an Internalizing factor, SCZ, BIP, and OCD on to a Thought Disorders factor, and ADHD, ASD, PTSD, and TS on to a Neurodevelopmental factor (Figure 1).

**Figure 1.**
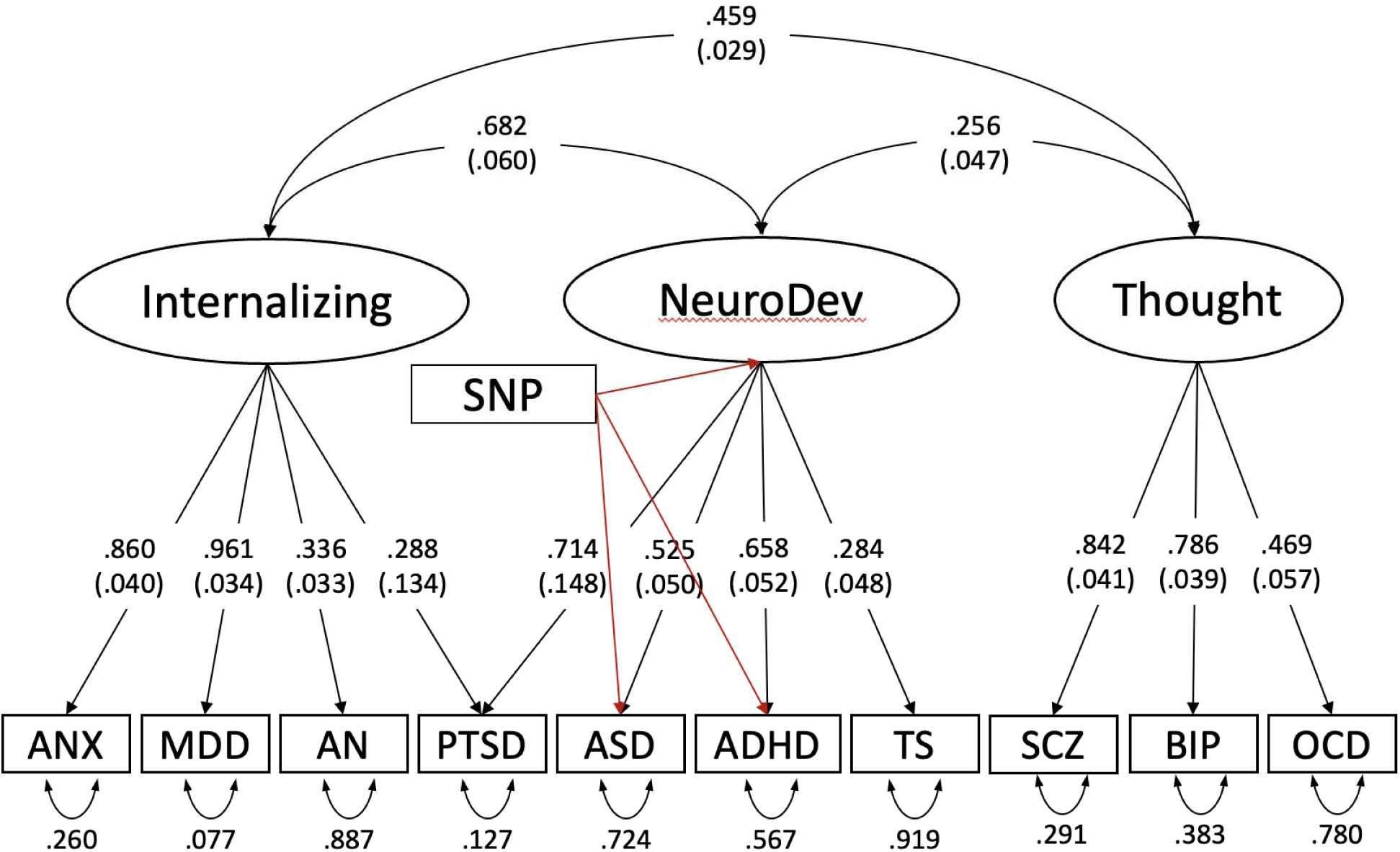
GenomicSEM model with 10 disorders, with specific SNP effects estimated for ASD, ADHD and the Neurodevelopmental latent factor

From this model, SNP effects were estimated for ADHD, ASD and the Neurodevelopmental latent factor in GenomicSEM. Manhattan and QQ plots for the GenomicSEM ADHD GWAS, GenomicSEM ASD GWAS and the Neurodevelopmental GWAS are provided in Figure S1. A list of the top SNP associations for each GWAS is provided in Tables S10-S12. Briefly, no SNP in the GenomicSEM ADHD GWAS reached threshold for genome-wide significance (*p* < 5 × 10^−8^). Only a single SNP – *SOX7* (rs10099100) – reached significance in the GenomicSEM ASD GWAS. Notably, rs10099100 was also one of five loci identified in the univariate ASD GWAS (15). In contrast, 19 independent loci were detected in the Neurodevelopmental GenomicSEM GWAS. The strongest SNP associations were near genes previously identified for neuronal development and neurodevelopmental disorders, including *PAPPA2* (rs147036913) (33), *SEMA6D* (rs35175834) (34), *PDE4B* (rs10789205) (35), and *CSMD1* (rs7830752) (36).

### Genetic Correlations

Figure 2A shows the genetic correlations of the PGS models with the 8 other GWAS summary statistics for psychiatric disorders. Figure 2B shows genetic correlations of the same PGS models with 17 GWAS of additional complex traits and diseases (also see Table S16 for effect sizes). Genetic correlations were robustly attenuated when comparing traditional PGS models to the GenomicSEM-tuned PGS models.

**Figure 2.**
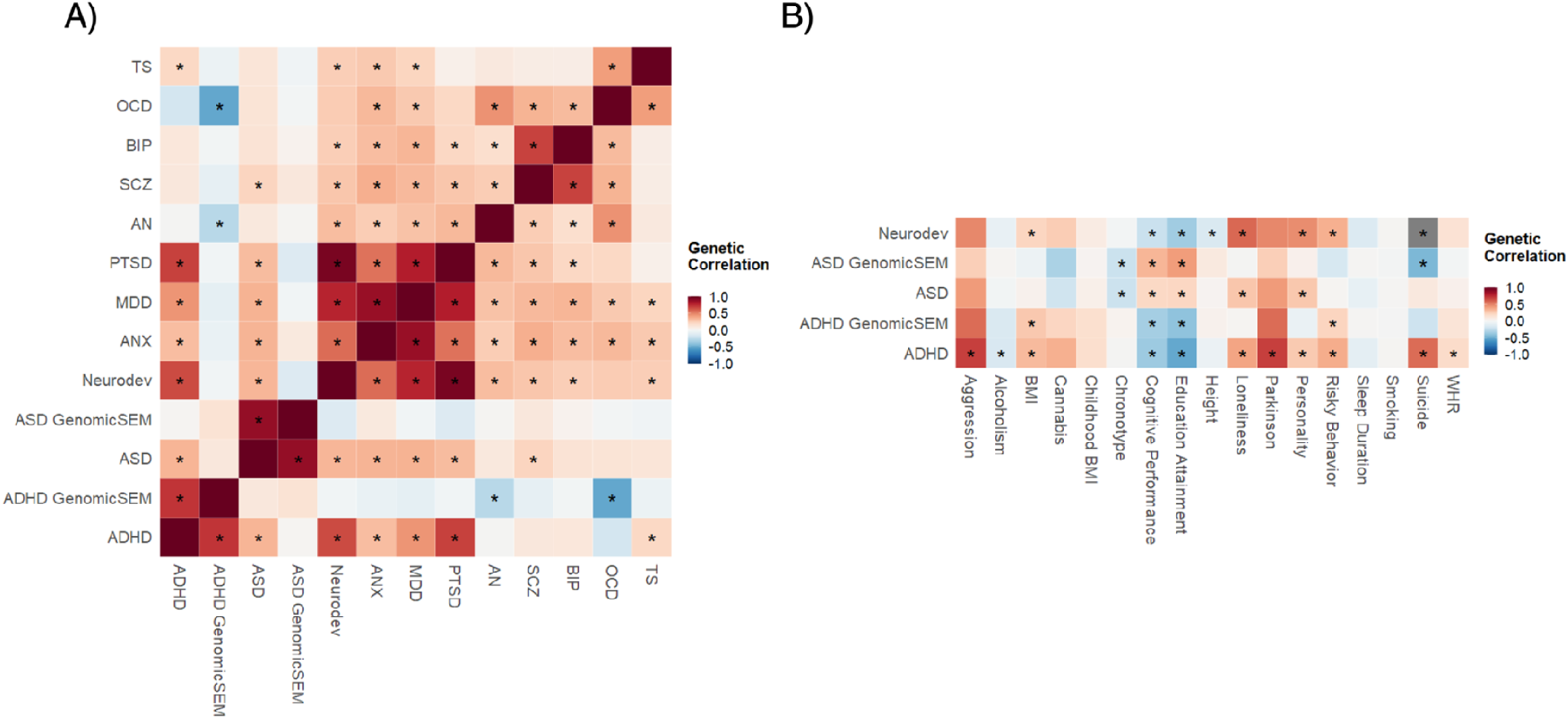
Genetic correlations. Panel A (left) shows genetic correlations between ADHD (traditional vs. GenomicSEM), ASD (traditional vs GenomicSEM), Neurodevelopmental PGS with 8 other psychiatric disorders that were included in the GenomicSEM model; Panel B (right) shows genetic correlations between ADHD (traditional and GenomicSEM), ASD (traditional vs. GenomicSEM), Neurodevelopmental PGS with 17 other complex traits and diseases (see Table S16 for details).

### PGS Associations with Focal (ADHD and ASD) Outcomes

#### PNC

First, we examined ADHD PGS associations with ADHD in a population-based sample in PNC (Table S13; Figure 3A). We expected all three PGS associations (traditional ADHD, GenomicSEM ADHD, and Neurodevelopmental) to be predictive of ADHD, with the GenomicSEM ADHD PGS having a weaker effect size compared to the traditional ADHD PGS. Indeed, all three PGS associations with ADHD were statistically significant: ADHD PGS (*B*=.332, *se*=.042, *p*.*fdr*<.001, *r*^2^=.012), GenomicSEM ADHD PGS (*B*=.207 *se*=.042, *p*.*fdr*<.001, *r*^2^=.005) and the Neurodevelopmental PGS (*B*=.147, *se*=.047, *p*.*fdr*<.001, *r*^2^=.002), with the GenomicSEM ADHD PGS showing an attenuated *r*^2^ relative to the ADHD PGS (Δ*r*^2^=.007).

**Figure 3.**
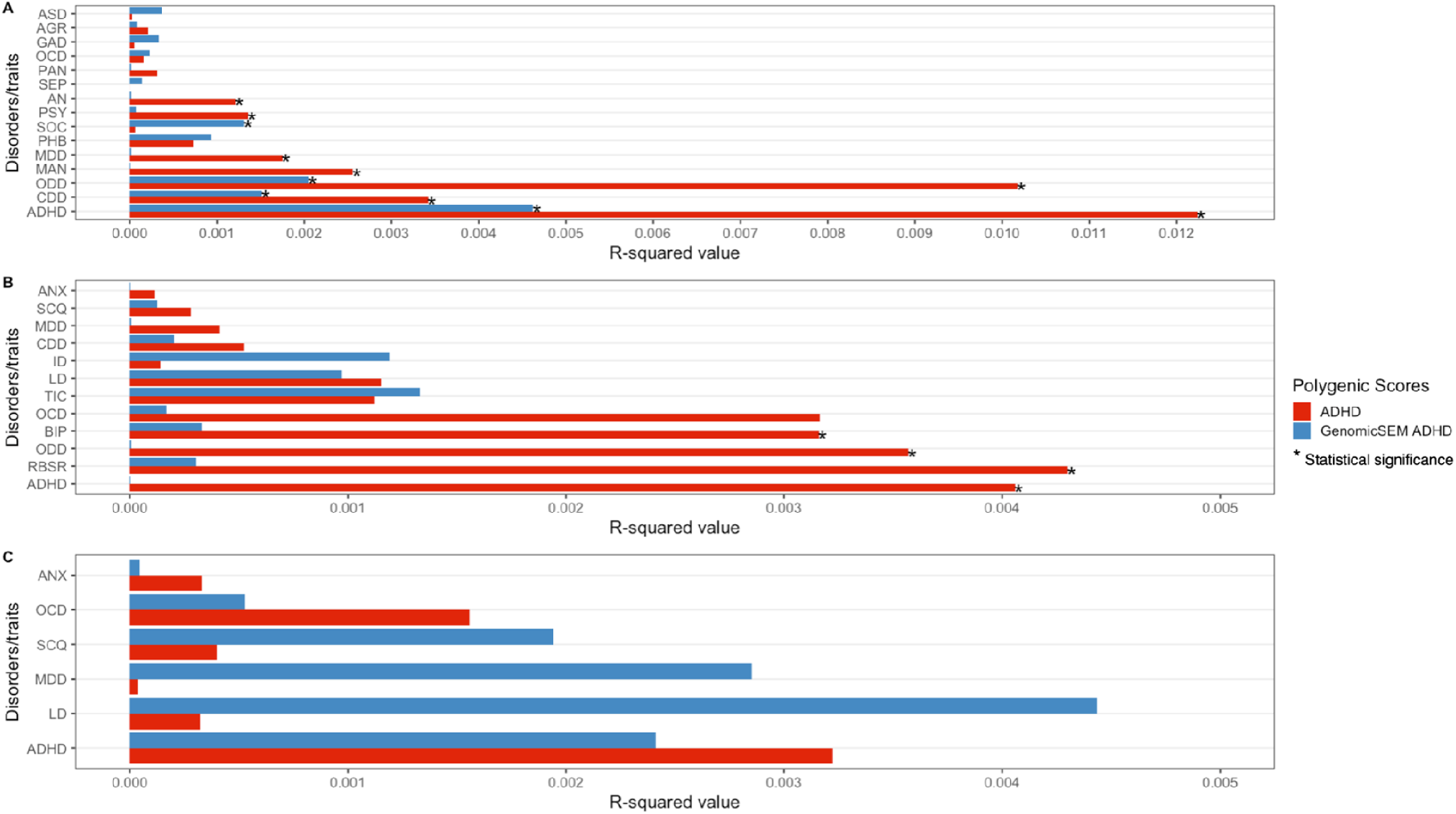
*r*^2^ of the ADHD PGS versus GenomicSEM ADHD PGS in predicting focal and non-focal outcomes across populations. Panel A (top) shows PGS associations with 15 psychiatric disorders in PNC; Panel B (middle) shows PGS associations with 12 psychiatric disorders or traits in SPARK probands; Panel C (bottom) shows PGS associations with 6 psychiatric disorders or traits in SPARK controls; *ASD: autism spectrum disorder; AGR: agoraphobia; GAD: generalized anxiety disorder; OCD: obsessive compulsive disorder; PAN: panic disorder; SEP: separation anxiety disorder; AN: anorexia; PSY: psychosis; SOC: social anxiety disorder; PHB: specific phobia; MDD: major depressive disorder; MAN: mania; ODD: oppositional defiant disorder; CDD: conduct disorder; ADHD: attention-deficit hyperactivity disorder; ANX: anxiety disorder; SCQ: social communication questionnaire; ID: intellectual disability; LD: learning disability; BIP: bipolar disorder; RBSR: repetitive behavior scale – revised*.

Then, we examined the traditional ASD PGS, GenomicSEM ASD PGS, and Neurodevelopmental PGS associations with ASD (Table S13; Figure 4A). Notably, both the traditional ASD and GenomicSEM ASD PGS associations were significant: ASD PGS (*B*=.359, *se*=.084, *p*.*fdr*<.001, *r*^2^=.005) and GenomicSEM ASD PGS (*B*=.301, *se*=.082, *p*.*fdr*<.001, *r*^2^=.004). The GenomicSEM ASD PGS showed an attenuated *r*^2^ (Δ*r*^2^=.001).

**Figure 4.**
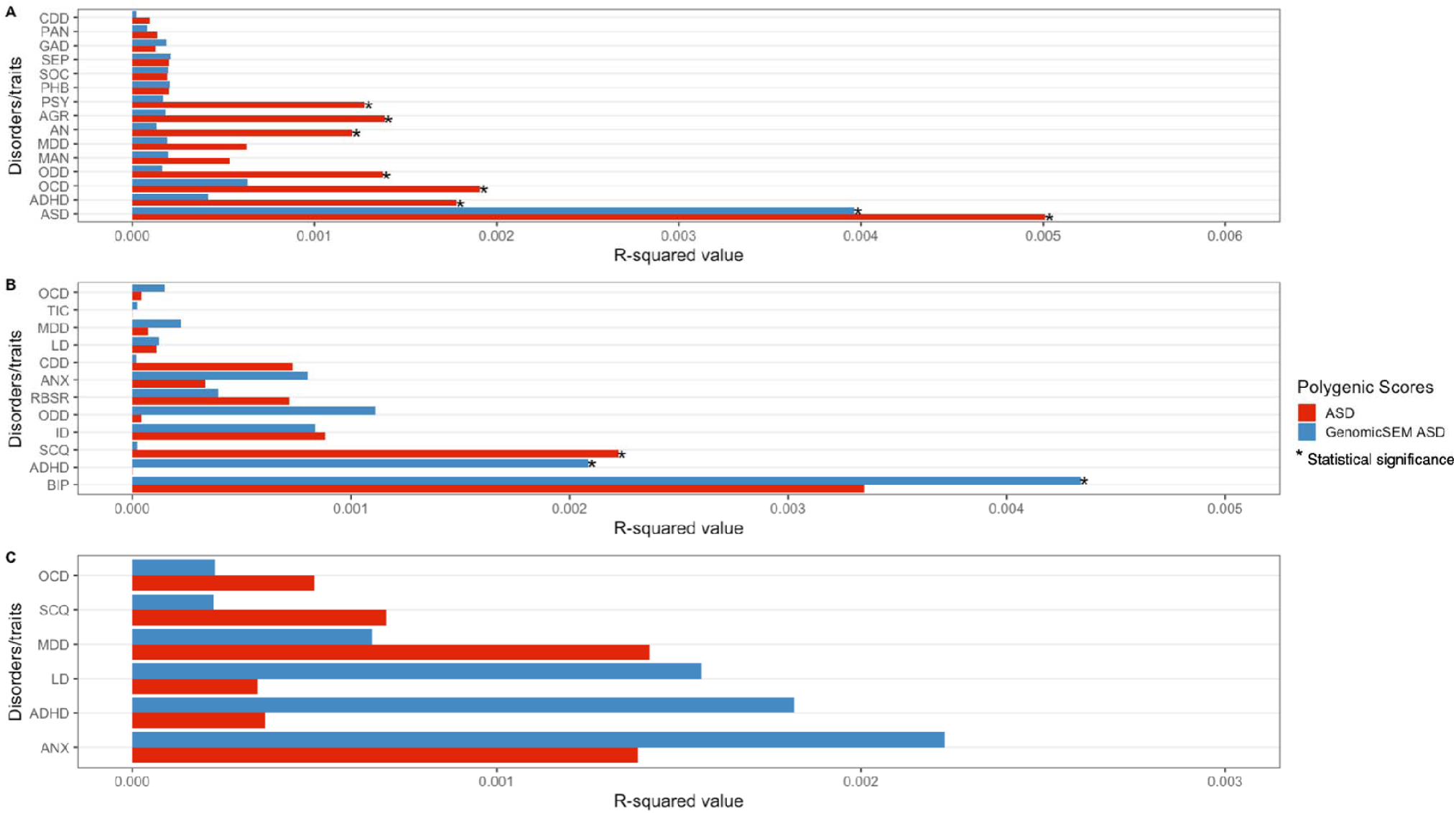
*r*^2^ of the ASD PGS versus GenomicSEM ASD PGS in predicting focal and non-focal outcomes across populations. Panel A (top) shows PGS associations with 15 psychiatric disorders in PNC; Panel B (middle) shows PGS associations with 12 psychiatric disorders or traits in SPARK probands; Panel C (bottom) shows PGS associations with 6 psychiatric disorders or traits in SPARK controls; *ASD: autism spectrum disorder; AGR: agoraphobia; GAD: generalized anxiety disorder; OCD: obsessive compulsive disorder; PAN: panic disorder; SEP: separation anxiety disorder; AN: anorexia; PSY: psychosis; SOC: social anxiety disorder; PHB: specific phobia; MDD: major depressive disorder; MAN: mania; ODD: oppositional defiant disorder; CDD: conduct disorder; ADHD: attention-deficit hyperactivity disorder; ANX: anxiety disorder; SCQ: social communication questionnaire; ID: intellectual disability; LD: learning disability; BIP: bipolar disorder; RBSR: repetitive behavior scale – revised*.

#### SPARK

Among SPARK probands, we expected the GenomicSEM ADHD PGS to show little to no prediction for ADHD given that this PGS was tuned to predict ADHD alone, rather than comorbidity (i.e., ASD + ADHD). Indeed, the traditional ADHD PGS was associated with ADHD (*B*=.045, *se*=.012, *p*.*fdr*=.003, *r*^*2*^=.004) (Table S14; Figure 3B) whereas the GenomicSEM ADHD PGS showed no association (*B*=-.001, *se*=.023, *p*.*fdr*=.992, *r*^*2*^=0). The Neurodevelopmental PGS was associated with ADHD status (*B*=.111, *se*=.034, *p*.*fdr*=.012, *r*^*2*^=.003). In other words, the GenomicSEM ADHD PGS demonstrated good discriminatory power within the ASD clinical sample, whereas the Neurodevelopmental PGS successfully predicted co-occurring status (i.e., ADHD + ASD).

We then examined the association of traditional ASD PGS, GenomicSEM ASD PGS, and the Neurodevelopmental PGS with RBSR and SCQ scores (i.e., ASD phenotypes) (Table S14, Figure 4B). The traditional ASD PGS was associated with the SCQ total score (*B*=.104, *se*=.036, *p*.*fdr*=.027, *r*^*2*^=.002), while no other PGS association was significant. Neither the traditional ADHD PGS, GenomicSEM ADHD PGS, nor the Neurodevelopmental PGS were associated with ADHD in SPARK controls (Table S15, Figure 3C). Similarly, the ASD PGS, GenomicSEM ASD PGS and the Neurodevelopmental PGS were not associated with ASD in SPARK controls (Table S15, Figure 4C).

### PGS Associations with Non-Focal Outcomes

#### PNC

Next, we tested the association of ADHD PGS and GenomicSEM ADHD PGS with non-focal outcomes in PNC (Table S13, Figure 3A). We expected ADHD PGS to demonstrate association signals across the psychiatric disorders, reflecting its genetic overlap with non-focal psychiatric disorders. GenomicSEM ADHD PGS should show attenuated or null prediction signals with non-focal psychiatric outcomes. For ease of interpretation, we report the number of significant (FDR corrected) associations that emerged, as individual effect sizes for all associations are reported in the supplemental tables.

The traditional ADHD PGS was not only associated with ADHD, but also for ODD, CDD, MDD, AN, MAN, and PSY (6 out of 14 outcomes, not including ADHD). In contrast, GenomicSEM ADHD PGS was only associated with ODD, CDD, and SOC (3 out of 14 outcomes). Associations between GenomicSEM ADHD PGS and ODD and CDD were attenuated compared to the models with ADHD PGS (Δ*r*^2^ = .008 and .001, respectively).

Next, we tested the discriminative performance of the traditional ASD PGS, GenomicSEM ASD PGS and the Neurodevelopmental PGS with non-focal (i.e., non-ASD) outcomes in PNC (Table S13, Figure 4A). The traditional ASD PGS was not only associated with ASD, but also for ADHD, ODD, AGR, AN, OCD, and PSY (6 out of 14 outcomes). Impressively, GenomicSEM ASD PGS was not associated with any non-focal outcomes (0 out of 14 outcomes), with the only signal emerging for ASD.

Not surprisingly, the Neurodevelopmental PGS was associated with half of the outcomes, including ODD, GAD, MDD, AN, MAN, OCD, and PSY (7 out of 14 outcomes) (Table S13).

#### SPARK

First, we examined the discriminatory performance of the traditional ADHD PGS and GenomicSEM ADHD PGS with non-focal outcomes in SPARK probands (Table S14, Figure 3B). ADHD PGS was associated with three non-ADHD outcomes – ODD, BIP, and the RBSR total score (3 out of 11 outcomes). GenomicSEM ADHD PGS was not associated with any of the non-ADHD outcome.

Next, we tested the discriminatory performance of the traditional ASD PGS and GenomicSEM ASD PGS with non-ASD outcomes. Again, as this subsample is entirely comprised of ASD cases, we did not expect the GenomicSEM ASD PGS to be free of non-focal associations as any residual GenomicSEM ASD PGS association may be reflective of ASD caseness. Indeed, the traditional ASD PGS was not associated with any non-focal outcome, whereas the GenomicSEM ASD PGS was associated with two non-ASD outcomes – ADHD and BIP (Table S14, Figure 4B).

In the SPARK controls, several outcomes exhibited low prevalence rates which resulted in model fitting errors (CDD, ODD, ID, BIP, TIC had lower than 1% prevalence). Additionally, the RBSR was not administered for control participants. ADHD PGS, ASD PGS, GenomicSEM ADHD PGS, GenomicSEM ASD PGS and the Neurodevelopmental PGS were not significantly associated with any of the disorders or traits (Table S15, Figures 3C and 4C).

## Discussion

Psychiatric PGS offer impressive predictive power but they suffer from poor discriminatory power. In this study, we used GenomicSEM to derive novel PGS for ADHD and ASD that provided superior discriminatory power for non-focal outcomes relative to traditionally computed PGS for these disorders. Moreover, a Neurodevelopmental PGS from GenomicSEM predicted genetically correlated multivariate phenotypes (i.e., co-occurring ADHD and ASD) across samples. This approach has clear clinical implications, as PGS that have better discriminative properties can be useful for psychiatric screening and diagnostic applications down the line.

Traditionally computed PGS for ADHD and ASD may be overgeneralized, as evidenced by their high degree of genetic correlation with other complex traits, as well as the inflated *r*^2^ for both focal and non-focal outcomes across two independent datasets. After we accounted for genetic covariation in GenomicSEM, the genetic correlations between ASD and ADHD with other psychiatric and complex traits were significantly attenuated. Additionally, no genome-wide significant variants were identified in the GenomicSEM ADHD GWAS, while only a single variant was detected in the GenomicSEM ASD GWAS. As a comparison, the ADHD GWAS conducted by the Psychiatric Genomics Consortium yielded 12 significant loci, whereas the ASD GWAS yielded five. Furthermore, the GenomicSEM GWAS for the Neurodevelopmental dimension (for which ADHD, ASD, PTSD, and TS all loaded on to) revealed 19 independent loci, with the top hits near genes that were identified for different disorders within this dimension. These findings suggest that many of the previously identified variants for ADHD (16) and ASD (15) are likely pleiotropic. Univariate psychiatric GWAS may not be suitable for identifying “genes for [psychiatric disorder]” as these GWAS often do not simultaneously assess for other, genetically correlated disorders in their samples. Statistical methods such as GenomicSEM may be suitable until psychiatric GWAS account for multiple phenotypes within the same discovery sample.

Next, we showed that GenomicSEM PGS for ADHD and ASD featured enhanced discriminatory power in outcome predictions, particularly in discriminating between phenotypes that are genetically correlated and challenging to differentiate clinically. Notably, the Neurodevelopmental PGS, which captures the genetic commonalities between ASD and ADHD via a higher order dimension, was predictive of ADHD within the ASD subgroup in SPARK. Thus, GenomicSEM was also useful for estimating pleiotropic genetic effects in terms of predicting psychiatric comorbidities (i.e., ADHD in an ASD subpopulation). Thus, one promising application of GenomicSEM is the uncovering of transdiagnostic etiological mechanisms.

While the superior discriminatory power of the GenomicSEM ADHD PGS held across both the non-clinical sample in PNC and the clinical ASD sample in SPARK, GenomicSEM ASD PGS was only more discriminative (compared to the traditional ASD PGS) in the non-clinical PNC sample, and not in the SPARK clinical sample. It should be noted that SPARK probands were entirely comprised of ASD cases such that GenomicSEM ASD PGS was highly unlikely to be free of non-focal associations in this subpopulation. The residual GenomicSEM ASD PGS associations in the ASD sample may be reflective of ASD caseness and the relatively high rate of comorbidity (specifically with ADHD and BIP) within the SPARK probands.

This study has a few limitations. First, we limited our analyses to individuals of European ancestry. As noted by Martin and colleagues (10), PGS perform poorly in non-European populations because genetic effect sizes are trained on predominantly European-ancestry discovery samples. Statistical methods are just now emerging that leverage GWAS from diverse populations to boost the statistical power of PGS models for non-European samples (37). However, these applications are still limited to non-psychiatric phenotypes. Another limitation is that the majority of PNC and SPARK individuals were children (i.e., individuals < age 18). Given the potential for confounding due to mixed-informant effects (i.e., adult self-report vs. parent-report of children’s psychiatric disorders), as well as the limited power to detect effects in the adult subsamples for PNC and SPARK, we limited our PGS association analyses to children. The issue of whether or not psychiatric PGS models can be made to be more developmentally sensitive is critical to address in future studies, as emerging studies have shown that psychiatric PGS may be better in predicting certain trajectories of psychopathology over others (38).

PGS may become a critical part of clinical assessments in the not-to-distant future (10). However, efforts to increase GWAS sample size for the sake of improving prediction *r*^2^ likely come at the cost of weaker specificity in the prediction signal as well, thus limiting its diagnostic utility. The current study demonstrates that statistical methods, like GenomicSEM, can be used to improve the discriminatory power of PGS from existing GWAS for single psychiatric outcomes by accounting for the genetic covariance of multiple psychiatric outcomes. This application could prove especially useful from a clinical perspective, as easy-to-administer and biologically informed assessment tools that are both sensitive and specific to the outcome are sorely needed.

## Data Availability

All data produced in the present work are contained in the manuscript.

## Acknowledgments

The authors were supported in part by a core grant to the Waisman Center from the *Eunice Kennedy Shriver* National Institute of Child Health and Human Development (P50HD105353).

