## Supplemental Figure 1 for "Enhancing the Discriminatory Power of ADHD and Autism Spectrum Disorder Polygenic Scores in Clinical and Non-Clinical Samples"

Supplemental Figures

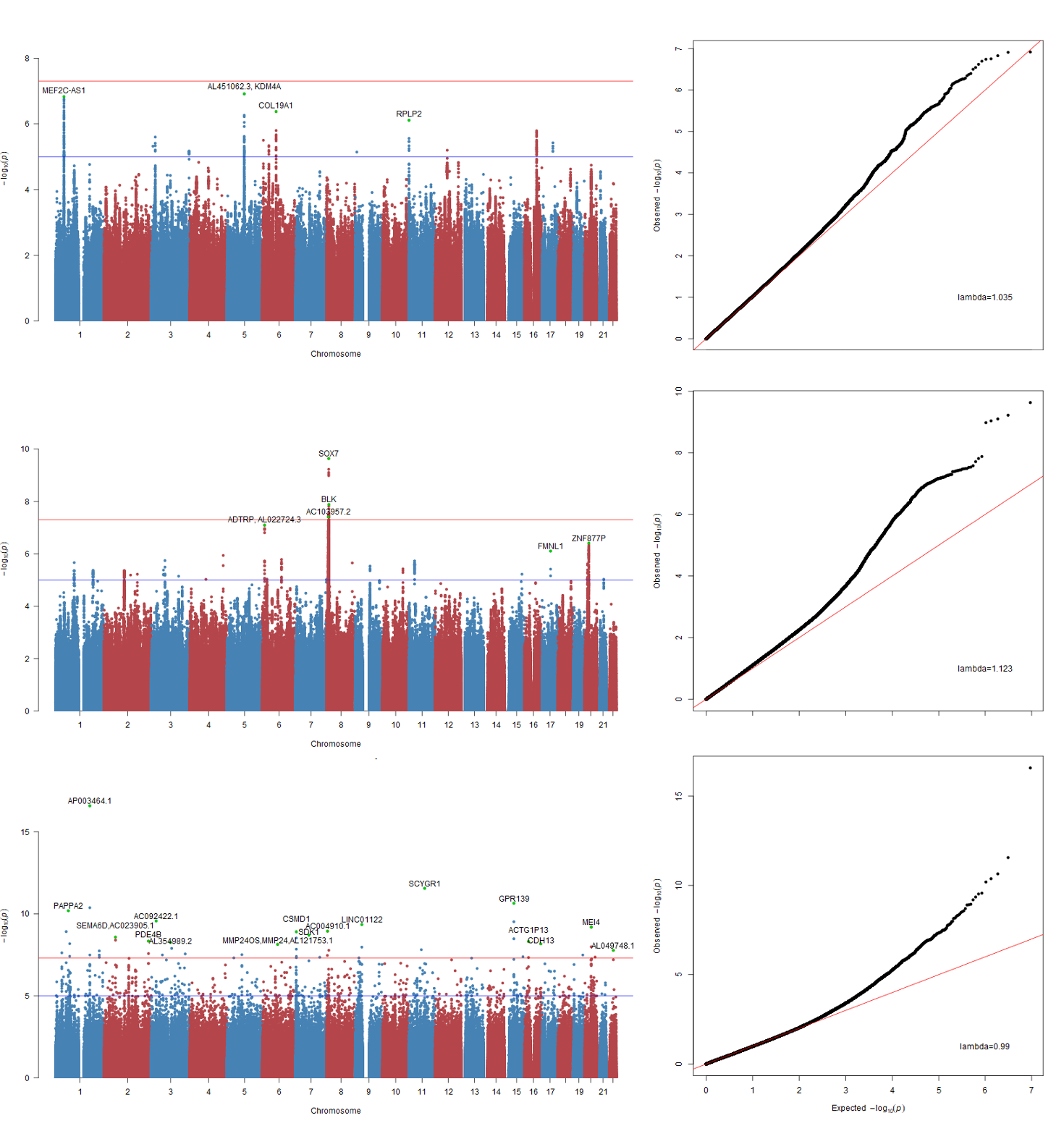

**Figure S1.** The Manhattan and QQ plots for GenomicSEM ADHD (top panels), GenomicSEM ASD (middle panels) and Neurodevelopmental GWAS (bottom panels).
